# Prevalence of Symptomatic Bacteriuria and Associated Risk Factors among Patients Attending Major Hospitals in Calabar, Nigeria

**DOI:** 10.1101/2023.09.26.23296138

**Authors:** Emmanuel E. Bassey, Maurice Mbah, Samuel S. Akpan, Edet E. Ikpi, Ambrose A. A. Alaribe

## Abstract

**Introduction:** Urinary tract infections (UTIs) are among the most encountered bacterial infection of humans and affect both male and female of all age groups, resulting in high mortality of infected patients if not properly managed.

**Hypothesis/Gap statement:** Several studies in different sub-Saharan Africa locations show variations in incidence of UTI as well as it’s causative organisms.

**Aim:** This study aimed to assess the prevalence, etiological agents, and factors associated with urinary tract infections among patients attending selected hospitals in Calabar metropolis, Nigeria.

**Methodology:** This was a cross-sectional study. Mid-stream urine samples collected from 240 patients with UTI were cultured using Cystine Lactose Electrolyte Deficient Agar (CLED). Data on socio-demographic, clinical symptoms and risk factors were obtained using structured questionnaire. Uropathogens were characterized using microbiological and biochemical tests and confirmed with API^®^ 20E and 20NE (Biomérieux) identification system. Pearson Chi-square was employed to check associations between categorical variables. *P*-value of <.05 was considered statistically significant.

**Results:** Out of the 240 urine samples collected, 13 were contaminated during collection, and 227 analyzed. Sixty-five (28.6%) patients had significant bacteriuria. Previous history of UTI (*P*=.000, CI=1.582-5.180), use of drugs without prescription (*P*=.000, CI=0.040-0.220), pregnancy (*P*=.001, CI=1.858-12.575) and history of urinary catheter (*P*=.031, CI=1.024-19.053) showed significant association with the occurrence of UTI. *Klebsiella pneumoniae* (23.1%) was the most predominant isolate, followed by Coagulase-negative Staphylococci (16.9%) and *Escherichia coli* (12.3%).

**Conclusion:** The study shows that routine UTI screening is beneficial for pregnant women, patients with difficult or painful urination, patients with previous episodes of UTI, catheterized patients and appropriate drugs administered for positive cases. In addition, self-administration of antibiotics should be avoided.

## INTRODUCTION

Urinary tract infection is any infection that occurs along the length of the urinary tract. It is characterized by the presence of bacteria in a supposedly sterile urinary tract, resulting in an increased bacterial load (often greater than 10^5^/mL) in urine sample ^[1]^. Urinary tract infections (UTIs) are widespread and affect a large proportion of the human population. Approximately, 150 million people are affected with UTI every year worldwide, with an estimated cost of U$5 billion each year in the United States ^[2]^. Although other microbial groups cause UTIs, the predominant organisms responsible for UTIs are members of the Enterobacteriaceae, with *E. coli* accounting for over 80% cases ^[3,4]^. Among the gram positive bacteria, *Staphylococcus aureus* and Coagulase-negative Staphylococci (CoNS) are mostly implicated in community acquired-UTIs. Fungi and viruses rarely cause UTI, however yeast, especially *Candida albicans*, are occasionally recovered from catheterized and/or immunocompromised patients ^[1]^. Other bacterial isolates implicated include *Klebseilla pneumoniae, Pseudomonas aeruginosa, Proteus mirabilis, Enterobacter clocae, Enterococcus feacalis* ^[2.3]^.

Factors for urinary tract infection vary with country and geographical location. Sex, personal hygiene, prostate problems, compromised immunity, diabetes, use of spermicidal contraception, and urinary catheterization are some of the factors for UTIs ^[4]^. Most cases of UTIs are seen in women due to their short urethra, proximity of the rectum to the urethra, change in vagina pH due to depletion of the commensal bacteria, hormonal imbalance occasioned by menstrual flow and/or pregnancy etc ^[5].^ Antibiotics resistance among uropathogens is a consistent problem making clinical management of the disease somewhat challenging ^[6]^. Our study showed that this influenced the prevalence rate of UTIs across different geographical areas. Several studies in different sub-Saharan Africa locations show variations in incidence of UTI as well as it’s causative organisms ^[7-10]^. Mwang’onde and Mchami ^[4]^ reported a median prevalence rate of 32.12% in sub-Saharan Africa and the commonest associated causative organism is *E. coli*.

Therefore, this study was conducted to assess the prevalence, etiological agents and factors associated with bacterial urinary tract infections, among patients attending hospitals in Calabar metropolis, Cross River State, Nigeria.

## MATERIALS AND METHODS

### Study Setting

This study was conducted in Calabar, the capital of Cross River State, located in Southern Nigeria. Calabar is administratively divided into Calabar Municipal and Calabar South Local Government Areas. It is located between latitudes 8°11’21” and 8°27’00” East of the Meridian and between latitudes 04°45’30” and 05°08’30” North of the Equator. Calabar metropolis has two major rivers: The Great Kwa River and Calabar River. The city has a total land area of four hundred and six (406) square kilometers ^[11]^.

According to 2006 Census Report, Calabar Metropolis had a total population of 375,196 ^[12]^. However, the rate of urbanization has greatly impacted on the population of the city ^[11]^. This population is primarily served by three hospitals: The University of Calabar Teaching Hospital (UCTH), Nigeria Naval Reference Hospital (NNRH) and General Hospital Calabar (GHC).

### Study Design/Study Subjects

A cross-sectional study was carried out between September and December, 2021 to assess the prevalence and associated risk factors of bacterial uropathogens among UTI patients attending major hospitals in Calabar, Cross River state, Nigeria. Inpatients and outpatients (≥ 5 years), clinically diagnosed with UTI were randomly recruited after informed consent was obtained. Patients who had taken antibiotics in the last seven days prior to the day of sample collection, <5 years old and those who willingly withdrew their consent were excluded from the study. Structured questionnaire was used to obtain information on socio-demographic (age, sex, educational status, occupation, and marital status). Female patients’ ≥12 years were screened for pregnancy. Sample size was calculated using a 95% confidence level, 5% precision and 19% prevalence of UTI ^[7]^. A total of 240 patients diagnosed with UTI participated in the study.

### Ethical Consideration

Ethical approval for this study was obtained from the Cross River State Health Research Ethics Committee (CRS-HREC) with **REC** No: CRSMOH/RP/2021/183. Data was collected after informed, voluntary, oral consent has been secured from each study subject. Confidentiality of subjects’ information was ensured. Positive results were reported to attending physician for appropriate treatment and management of the patients.

### Urine Collection and Analysis

Clean catch mid-stream urine was collected into sterile universal containers with each bottle labeled with the patient’s identity number and the assigned hospital code. Samples were subsequently conveyed for evaluation and microbiological analysis. Uropathogens were isolated using semi-quantitative culture techniques previously described by Kass ^[13]^. Each urine sample was inoculated on Cystine Lactose Electrolyte Deficient Agar (CLED) plates using calibrated loop (0.002 mL) and thereafter incubated aerobically at 37°C for 18-24 hours. Colony forming units (CFU) were determined to check for significant bacteriuria. Urine samples with colonies ≥10^5^CFU/mL were considered significant. Distinctive colonies from significant cultures were sub-cultured into fresh agar plates.

### Phenotypic Characterization/Identification

The isolated uropathogens were identified using colonial morphology, growth characteristics, Gram stain, conventional biochemical tests ^[14]^ and confirmed with commercially prepared biochemical test kits: API 20E and API 20NE (Biomérieux). The results obtained were entered into Analytical Profile Index (API) software for identification of the isolates.

### Screening for Pregnancy

Pregnancy was screened using a qualitative human chorionic gonadotropin (hCG) urine test ^[10]^. In this test, LabACON^®^ (Hangzhou Biotest Biotec Co., Ltd) pregnancy rapid test strip was used to detect hCG hormones in patient’s urine samples. The test strip was dipped into a bottle of freshly collected urine and the result read within 3 minutes. For positive result, two lines colour bands were observed. For negative result, only one colour band was observed.

### Macroscopic and Microscopic Examination of Urine

Prior to microscopic examination, each specimen was checked macroscopically for colour and transparency and the results recorded. Each sample (5 mL) of urine was centrifuged for ten minutes at 3000 rpm. The supernatant was examined with x10 objective lens and then x40 with the condenser iris closed. Samples with leukocytes ≥ 10 per high power field (HPF) (x40 objective) were considered pyuria ^[14]^.

### Statistical Analysis

Data derived from questionnaire and microbiological survey were analyzed using Statistical Package for Social Sciences (SPSS) version 20.0. Descriptive statistics, such as frequencies, percentages, means and standard deviations, were used. Pearson Chi-square was employed to check associations between categorical variables. *P*-value of <.05 was considered statistically significant.

## RESULTS

### Socio-Demographic Data of Participants

Two hundred and forty (240) participants who met the inclusion criteria were enrolled in the study. Among this numbers, ninety-eight (98) participants were recruited from NNRH, 78 from GHC and 62 from UCTH. Thirteen (13) out of 240 MSU samples collected were contaminated due to inappropriate samples collection measures. More females 59.5% (135/227) participated in the study compared to males 40.5% (92/227). The age of the study participants ranged from 5 to 75 years with the mean age of 34.1± 0.8 (Table 1). A proportion of 39.6% (90/227) were in the age range 30-45 years and 49.3% (112/227) were single. Most participants (56.4%, 128/227) acquired tertiary educational qualification while 44.9% (102/227) of the respondents were students (Table 1).

**TABLE 1:** Socio-Demographic Data, Clinical Symptoms and Risk Factors for UTI.

| Characteristics | Number (%) | NNRH n(%) | UCTH n(%) | GHC n(%) |
| --- | --- | --- | --- | --- |
| **Age** |  |  |  |  |
| 5-18 | 15 (6.6) | 9 (9.6) | 0 (0.0) | 6 (8.5) |
| 19-29 | 82 (36.1) | 31 (33.0) | 17 (27.4) | 34 (47.9) |
| 30-45 | 90 (39.6) | 41 (43.6) | 28 (45.2) | 21 (29.6) |
| Above 45 | 40 (17.6) | 13 (13.8) | 17 (27.4) | 10 (14.1) |
| **Sex** |  |  |  |  |
| Male | 92 (40.5) | 37 (39.4) | 29 (46.8) | 26 (36.6) |
| Female | 135 (59.5) | 57 (60.6) | 33 (53.2) | 45 (63.4) |
| **Marital Status** |  |  |  |  |
| Single | 112 (49.3) | 43 (45.7) | 24 (38.7) | 45 (63.4) |
| Married | 110 (48.5) | 47 (50.0) | 37 (59.7) | 26 (36.6) |
| Divorced | 3 (1.3) | 3 (3.2) | 0 (0.0) | 0 (0.0) |
| Widow | 0 (0.0) | 0 (0.0) | 0 (0.0) | 0 (0.0) |
| Widower | 2 (0.9) | 1 (1.1) | 1 (1.6) | 0 (0.0) |
| **Educational Qualification** |  |  |  |  |
| No formal education | 9 (4.0) | 2 (2.1) | 5 (8.1) | 2 (2.8) |
| Primary | 25 (11.0) | 10 (10.6) | 8 (12.9) | 7 (9.9) |
| Secondary | 65 (28.6) | 33 (35.1) | 15 (24.2) | 17 (23.9) |
| Tertiary | 128 (56.4) | 49 (52.1) | 34 (54.8) | 45 (63.4) |
| **Major Occupation** |  |  |  |  |
| Farming | 16 (7.0) | 3 (3.2) | 9 (14.5) | 4 (5.6) |
| Fishing | 0 (0.0) | 0 (0.0) | 0 (0.0) | 0 (0.0) |
| Trading | 46 (20.3) | 20 (21.3) | 15 (24.2) | 11 (15.5) |
| Student | 102 (44.9) | 38 (40.4) | 22 (35.5) | 42 (59.2) |
| Civil servant | 56 (24.7) | 33 (35.1) | 9 (14.5) | 14 (19.7) |
| Artisan | 7 (3.1) | 0 (0.0) | 7 (11.3) | 0 (0.0) |
| **Clinical Symptoms** |  |  |  |  |
| Urine urgency | 12 (5.3) | 4 (4.3) | 6 (9.7) | 2 (2.8) |
| Dysuria | 46 (20.3) | 17 (18.1) | 16 (25.8) | 13 (18.3) |
| Fever | 54 (23.8) | 25 (26.6) | 8 (12.9) | 21 (29.6) |
| Flank or supra-pubic pain | 50 (22.0) | 23 (24.5) | 9 (14.5) | 18 (25.4) |
| Frequency of urination | 59 (26.0) | 16 (17.0) | 25 (40.3) | 19 (26.8) |
| **Risk Factors** |  |  |  |  |
| Pregnancy status | 22 (16.3) | 9 (15.8) | 5 (15.2) | 8 (17.8) |
| Use of drug without prescription | 63 (15.9) | 24 (25.5) | 15 (24.2) | 24 (33.8) |
| Previous history of UTI | 85 (37.4) | 44 (46.8) | 18 (29.0) | 23 (32.4) |
| History of catheterization | 8 (3.8) | 2 (2.1) | 5 (8.1) | 1 (1.4) |
| Family history of UTI | 121 (53.3) | 48 (51.1) | 33 (53.2) | 42 (59.2) |
| Diabetes | 3 (1.3) | 0 (0.0) | 2 (3.2) | 1 (1.4) |
| Use of contraceptive | 27 (20.0) | 7 (12.3) | 6 (18.2) | 14 (31.1) |

NNRH = Nigeria Navy Reference Hospital Calabar
UCTH = University of Calabar Teaching Hospital
GHC = General Hospital Calabar

### Clinical Symptoms and Risk Factors of UTI

Based on responses to the structured questionnaire, clinical symptoms and potential underlying risk factors for UTI were evaluated. Clinical symptoms and risk factor for UTI examined among participants is depicted in (Table 1). The clinical symptoms among the study subjects were 26.0% (59/227), 23.8% (54/227), 22.0% (50/227), 20.3% (46/227) and 5.3% (12/227) for frequency of urination, fever, flank or supra-pubic pain, dysuria and urine urgency, respectively. Of all considered risk factors, 53.3% (121/227) of the respondents had family relative with history of UTI. Eighty-five (37.4%) had recurrent (i.e. previous history of UTI) 15.9% (63/227) used drug without prescription, 3.5% (8/227) had history of urinary catheter while 1.3% (3/227) patients were diabetic. Out of 135 female participants, 20.0% used contraceptives and 16.3% were pregnant (Table 1).

### Prevalence of UTI Among Participants

This study revealed that, out of 227 samples analyzed, 65 showed significant bacterial growth, giving an overall prevalence of 28.6% (Table 2). However, in NNRH, 94 urine samples were analyzed, 31 showed significant bacterial growth, giving a UTI prevalence of 33.0%. Results from UCTH showed that 15 (24.2%) out of 62 samples analyzed were positive for UTI. Seventy-one urine samples were analyzed from patients in GHC, significant bacteriuira was found in 26.8% (19/71) of the patients (Table 2).

**Table 2:** Socio-Demographic Data of Participants and their Associations with UTIs.

| **PREVALENCE** | **NNRH (N = 94)**<br>**n(%)** | **UCTH (N = 62)**<br>**n(%)** | **GHC (N = 71)**<br>**n(%)** | **TOTAL (N = 227)** | **P-value** |
| --- | --- | --- | --- | --- | --- |
| Significant bacteriuria | 31 (33.0%) | 15 (24.2%) | 19 (26.8%) | 65 (28.6%) |  |
| No Significant bacteriuria n(%) | 63 (67.0%) | 47 (75.8%) | 52 (73.2%) | 162 (71.4%) |  |
| **Socio-Demographic Data** | **Sig. bacteriuria n(%)** | **Sig. bacteriuria n(%)** | **Sig. bacteriuria n(%)** |  |  |
| **Age** |  |  |  |  |  |
| 5-18 | 4 (12.9) | 0 (0.0) | 0(0.0) | 4 (6.2) | .946 |
| 19-29 | 9 (29.0) | 4 (26.4) | 12(63.2) | 25 (38.5) |  |
| 30-45 | 13 (41.9) | 6 (40.0) | 5(26.3) | 24 (37.0) |  |
| Above 45 | 5 (16.1) | 5 (33.3) | 2(10.5) | 12 (18.5) |  |
| **Sex** |  |  |  |  |  |
| Male | 11 (35.5) | 11 (73.3) | 4 (21.1) | 26 (40.0) | .918 |
| Female | 20 (64.5) | 4 (26.7) | 15 (78.9) | 39 (60.0) |  |
| **Marital status** |  |  |  |  |  |
| Single | 13 (41.9) | 3 (20.0) | 11 (57.9) | 27 (41.5) | .347 |
| Married | 17 (54.8) | 12 (80.0) | 8 (42.1) | 37 (57.0) |  |
| Divorced | 1 (3.2) | 0 (0.0) | 0 (0.0) | 1 (1.5) |  |
| Widow | 0 (0.0) | 0 (0.0) | 0 (0.0) | 0 (0.0) |  |
| Widower | 0 (0.0) | 0 (0.0) | 0 (0.0) | 0 (0.0) |  |
| **Educational qualification** |  |  |  |  |  |
| No formal education | 0 (0.0) | 1 (6.7) | 0(0.0) | 1 (1.5) | .508 |
| Primary | 3 (9.7) | 3 (20.0) | 1(5.3) | 7 (10.8) |  |
| Secondary | 12 (38.7) | 6 (40.0) | 4(21.1) | 22 (33.8) |  |
| Tertiary | 16 (51.6) | 5 (33.3) | 14(73.7) | 35 (53.8) |  |
| **Major occupation** |  |  |  |  |  |
| Farming | 0 (0.0) | 1 (6.7) | 0(0.0) | 1 (1.5) | .003* |
| Fishing | 0 (0.0) | 0 (0.0) | 0(0.0) | 0 (0.0) |  |
| Trading | 7 (22.6) | 2 (13.3) | 2 (10.5) | 11 (17.0) |  |
| Student | 11 (35.5) | 3 (20.0) | 11 (57.9) | 25 (38.5) |  |
| Civil servant | 13 (41.9) | 4 (26.7) | 6 (31.6) | 23 (35.4) |  |
| Artisan | 0 (0.0) | 5 (33.3) | 0(0.0) | 5 (7.7) |  |

\*Statistically significant at $P < .05$

### Associated Socio-Demographic Data with UTI

Prevalence of UTI was higher in females (60%, 39/65) than in males (40%, 26/65). Prevalence of UTI was higher among patients in the age group 19-29 years (38.5%). Prevalence of UTI in relation to marital status showed that majority (57.0%, 37/65) were married. Among those diagnosed with UTI, patients with tertiary educational qualification were the most encountered (53.8%, 35/65). Based on subjects’ occupational status, students had the highest prevalence of UTI (38.5%, 25/65). However, age, sex, marital status and educational qualification showed no significant (*P*>.05) association with the occurrence of UTI. Participants occupation showed statistical significant (*P*=.003) association with UTI (Table 2).

### Associated Clinical Symptoms & Risk Factors with UTI

The results revealed that of all assessed clinical symptoms, dysuria (*P*=.033, CI=1.051-4.059) showed statistical association with UTI. Pearson Chi-Square showed that urine urgency (*P*=.305, CI=0.564-6.039), fever (*P*=.124, CI=0.270-1.177), flank or supra-pubic pain (*P*=.412, CI=0.358-1.524) and frequency of urination (*P*=.481, CI=0.134-1.187) were not statistically associated with UTI.

Association between the risk factors and the occurrence of UTI showed that 57.0% (37/65) of the respondents had family history of UTI. Thirty-six (55.4%) had recurrent (i.e. previous UTI history) cases, 27.7% (18/65) used drug without prescription, 7.7% (5/65) had history of urinary catheter while 3.1% (2/65) patients were diabetic. Out of 39 females positive for UTI, 25.6% used contraceptives and 33.3% were pregnant (Table 3). The findings revealed that history of UTI (*P*=.000, CI=1.582-5.180), use of drugs without prescription (*P*=.000, CI=0.040-0.220), pregnancy (*P*=.001, CI=1.858-12.575) and history of urinary catheter (*P*=.031, CI=1.024-19.053) showed significant association with the occurrence of UTI. However, other factors like family history of UTI, diabetes and use of contraceptive showed no significant relationship with the occurrence UTI (*P*>.05) (Table 3).

**TABLE 3:** Clinical Symptoms, risk Factors and their Associations with UTIs.

| PREVALENCE |  | NNRH (N = 94) | UCTH (N = 62) | GHC (N = 71) | TOTAL | p-value, (95% CI) |
| --- | --- | --- | --- | --- | --- | --- |
|  |  | n(%) | n(%) | n(%) | (N = 227) |  |
| **Clinical symptoms** |  | Sig. bacteriuria | Sig. bacteriuria | Sig. bacteriuria |  |  |
|  |  | n(%) | n(%) | n(%) |  |  |
| Urine urgency | Yes | 3 (9.7) | 0(0.0) | 2(10.5) | 5 (7.7) | .305, (0.564-6.039) |
|  | No | 28 (90.3) | 15(100) | 17(89.5) | 60 (92.3) |  |
| Dysuria | Yes | 7 (22.6) | 5(33.3) | 7(36.8) | 19 (29.2) | **.033\***, (1.051-4.059) |
|  | No | 24 (77.4) | 10(66.7) | 12(63.2) | 46 (70.8) |  |
| Fever | Yes | 4 (12.9) | 1(6.7) | 6(31.6) | 11 (17.0) | .124, (0.270-1.177) |
|  | No | 27 (87.1) | 14(93.3) | 13(68.4) | 54 (83.1) |  |
| Flank or supra-pubic pain | Yes | 7 (22.6) | 2(13.3) | 3(15.8) | 12 (18.5) | .412, (0.358-1.524) |
|  | No | 24 (77.4) | 13(86.7) | 16(84.2) | 53 (81.5) |  |
| Frequency of urination | Yes | 4 (12.9) | 9 (60.0) | 7 (36.8) | 19 (29.2) | .481, (0.134-1.187) |
|  | No | 27 (87.1) | 6( 40.0) | 12 (63.2) | 46 (70.8) |  |
| **Risk factors** |  |  |  |  |  |  |
| Pregnancy status | Yes | 5 (25.0) | 3(75.0) | 5 (33.3) | 13 (33.3) | **.001\***, (1.858-12.575) |
|  | No | 15 (75.0) | 1(25.0) | 10 (66.7) | 26 (66.7) |  |
| Use of drug without prescription | Yes | 7 (22.6) | 5(33.3) | 6 (31.6) | 18 (27.7) | **.000\***, (0.040-0.220) |
|  | No | 24 (77.4) | 10(66.7) | 13 (68.4) | 47 (72.3) |  |
| Previous history of UTI | Yes | 17 (54.8) | 11(73.3) | 8(42.1) | 36 (55.4) | **.000\***, (1.582-5.180) |
|  | No | 14 (45.2) | 4(26.7) | 11(57.9) | 29 (44.6) |  |
| History of catheterization | Yes | 1 (3.2) | 3(20.0) | 1(5.3) | 5 (7.7) | **.031\***, (1.024-19.053) |
|  | No | 30 (96.8) | 12(80.0) | 18(94.7) | 60 (92.3) |  |
| Family history of UTI | Yes | 16 (51.6) | 9(60.0) | 14(73.7) | 37 (57.0) | .489, (0.687-2.191) |
|  | No | 15 (48.4) | 6(40.0) | 5(26.3) | 28 (43.1) |  |
| Diabetes | Yes | 0 (0.0) | 2(13.3) | 0(0.0) | 2 (3.1) | .142, (0.455-57.368) |
|  | No | 31 (100) | 13(86.7) | 19(100) | 63 (97.0) |  |
| Use of contraceptive | Yes | 2 (10.0) | 2(50.0) | 6(40.0) | 10 (25.6) | .296, (0.658-3.900) |
|  | No | 18 (90.0) | 2(50.0) | 9(60.0) | 29 (74.4) |  |

\*Statistically significant at $P < .05$

### Prevalence of Uropathogens from the Study

Ten different bacterial species were isolated from the samples. Gram negative uropathogens constituted 80.0% (52/65) while 20% (13/65) were Gram positive cocci. The most predominant isolated bacteria were *Klebsiella pneumoniae* (23.1%). This was followed by Coagulase-negative Staphylococci (CoNS) (16.9%), *Escherichia coli* (12.3%), *Enterobacter clocae* (10.8%), *Citrobacter freundii* (9.2%), *Proteus mirabilis, Serratia marcescens* with (7.7%) each, *Pseudomonas aeruginisa, Cronobacter* sp, *Enterococcus* sp with (3.1%) each, while *Citrobacter koseri* and *Pseudomonas luteola* (1.5%) were the least isolated uropathogens (Table 4).

**TABLE 4:**
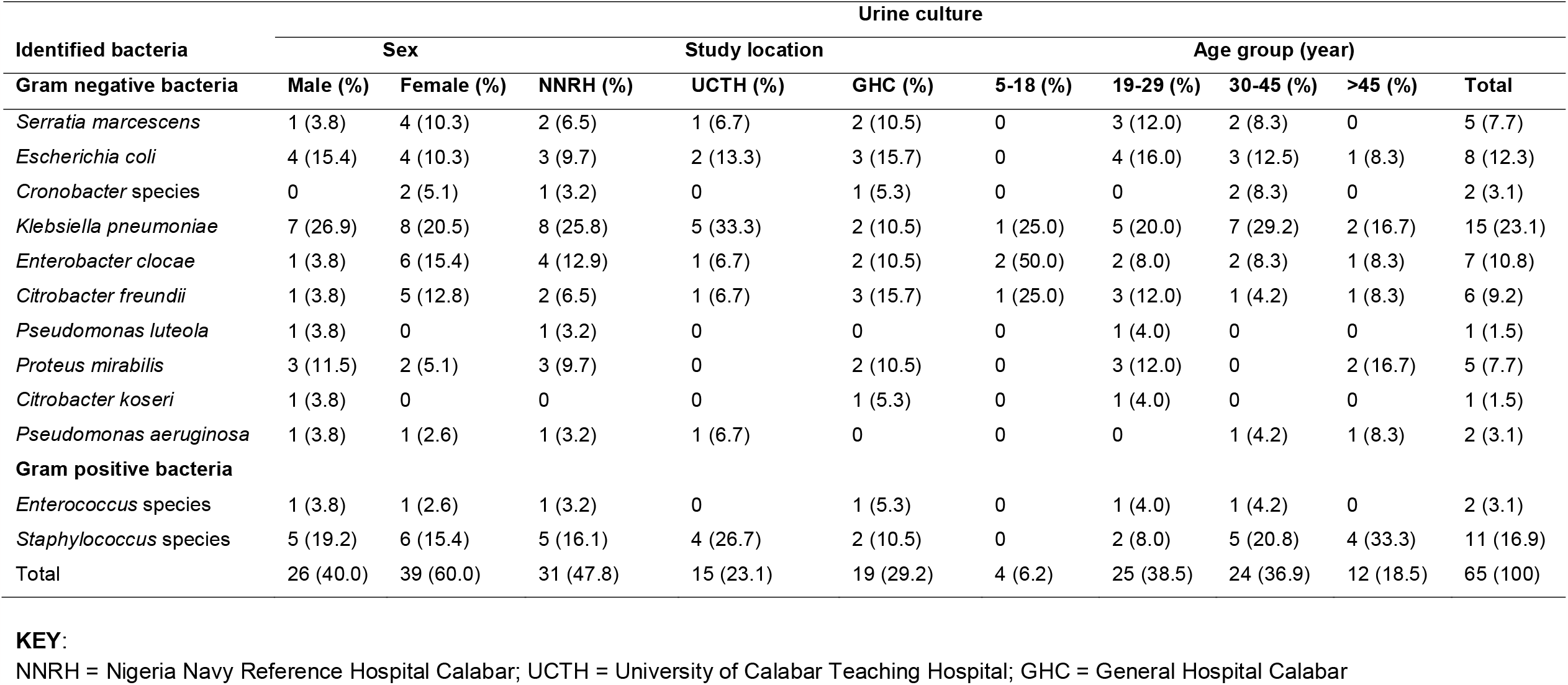
Prevalence of uropathogens isolated from patients by gender, study site, and age group.

### Prevalence of Uropathogens by Study Sites

The highest prevalence uropathogens was recovered from patients in NNRH (47.8%), followed by GHC (29.2%) while 23.1% of the isolated uropathogens were from UCTH. The most encountered uropathogen in NNRH was *K. pneumoniae* (25.8%). This was followed by CoNS (16.1%), *E. clocae* (12.9%), *E. coli* and *P. mirabilis* had 9.7% each, *S. marcescens* and *C. freundii* were recovered at the rate of 6.5% each, while the least isolated uropathogens were *Cronobacter* sp, *P. luteola, P. aeruginosa* and *Enterococcus* sp at 3.2%, 3.2%, 3.2% and 3.2% respectively (Table 4). Similarly, *K. pneumoniae* (33.3%) was the most isolated uropathogen in UCTH. Others were CoNS (26.7%), *E. coli* (13.3%), *S. marcescens* (6.7%), *E. clocae* (6.7%), *C. freundii* (6.7%) and *P aeruginosa* (6.7%) (Table 4). On the contrary, *E. coli* and *C. freundii* were the most occurrence uropathogens in GHC with prevalence rate of 15.7% each, followed by *S. marcescens* (10.5%), *K. pneumoniae* (10.5%), *E. clocae* (10.5%), *P. mirabilis* (10.5%), CoNS (10.5%), and the least occurred uropathogens were *Cronobacter* sp, *C. koseri* and *Enterococcus* sp at 5.3% each (Table 4).

### Prevalence of Uropathogens by age Group

The highest prevalence of uropathogens was observed in patients in the age group 19-29 years (38.5%) followed by 30-35 years (36.9%), >45 years (18.5%) while the least bacterial uropathogens was recovered from age group 5-18 years at 6.2% (Table 4). *K. pneumoniae* were the most isolated uropathogens in the age group 19-29 years and 30-45 years. In the age group 5-18 years, predominant uropathogens was *E. coli* whereas CoNS was common in the age group 45 years. Statistical analysis revealed that there was no significant association between age group and the occurrence of UTI, *P*= .946, χ^2^ = 0.373 (Table 2).

### Prevalence of Uropathogens by Gender

The study showed that out of 65 recovered uropathogens, 60.0% were from females while 40.0% were from males. *K. pneumoniae* and CoNS were the most encountered uropathogens in males whereas *K. pneumoniae*, CoNS and *E. clocae* were mostly seen in females (Table 4). Statistical analysis revealed that there was no significant relationship between gender and the prevalence of UTI, *P*= .918, χ^2^= 0 .011 (Table 2).

## DISCUSSION

In this study, 65 out 227 analyzed samples yielded significant growth giving an overall prevalence of 28.6% among the study participants. This is similar to 29.0% reported in Ismaila City, Egypt ^[15]^. The prevalence of UTI recorded in this study is somewhat similar to 31.3% reported from among symptomatic UTI patients in Enugu, Nigeria ^[16]^. Somewhere outside the shore of Nigeria, Odoki *et al*. ^[10]^ reported 32.2% in Bushenyi District, Uganda, which is comparable to the present study. The prevalence of UTI in the present study was slightly higher than the ones previously reported in the same study area ^[7,17]^. Other researchers have reported prevalence rates of UTI higher or slightly lower when compared to the current study ^[18-20]^. The observed difference in prevalence of UTI could be affected by personal hygiene, study population, socio-economic status, immune status of the study participants and geographical variation.

Interestingly, the highest prevalence (33%) of UTI was recorded among patients who visited the NNRH when compared to other study sites (Table 2). The higher prevalence of UTI among these patients could be attributed to the status of NNRH as reference hospital where advanced level of healthcare services and diagnoses are conducted. This observation is in consonance with the work of Karikari *et al*. ^[9]^ in Ghana, who recorded high incidence of UTI in a referral hospital.

The age group 19-29 years had the highest prevalence of UTI (38.5%). However, 30-45 years had the highest prevalence of UTI in UCTH and NNRH while the age group 19-29 years recorded the highest incidence of UTI in GHC. This finding is in consonance with Ndako *et al*. ^[19]^ in South-West Nigeria, Abdu *et al*. ^[21]^ in Maiduguri, Nigeria, and Mokube *et al*. ^[8]^ in Cameroon. In contrast, Obirikorang *et al*. ^[22]^ reported highest prevalence of UTI in age group 30-34 years in Kumasi, Ghana, Marami *et al*. ^[23]^ in age group 35-44 years in Ethiopia. However, age was not found significantly (*P*=.946) associated with the occurrence of UTI. This variation in prevalence of UTI in age group might be due to the target population under study, previous untreated history, family history, underlying medical condition, sexual activity, and educational level, among other factors ^[5,24]^.

In this study, UTI was higher in females (60%, 39/65) than in males (40%, 26/65). This finding is consistence with previous studies supporting that women are more vulnerable to contracting UTI due to their short urethra and its proximity to the anal opening ^[25,26]^. Other factors which may contribute to high incidence of UTI in women are physiological changes in women which deplete the virginal flora, unauthorized administration of contraceptives, pregnancy, family history, etc ^[4]^.

Prevalence of UTI was higher in married subjects (57%) compared to single (41.5%) (Table 2), but in GHC, single participants had the highest prevalence rate of UTI (57.9%). This is in agreement with Ezugwu *et al*. ^[27]^ but contradict Wanja *et al*. ^[28]^ in Kenya, who reported highest prevalence of UTIs among widowed (57.1%). The highest prevalence of UTI among single participants in GHC may be attributed to the proximity of UNICAL to this site. Multiple sex partners and unfaithfulness to one sex partner have been found as an indicator of UTI among university students ^[29]^.

Participants with tertiary educational level had the highest prevalence of UTI (53.8%, 35/65). But in UCTH, subjects who attended secondary school had the highest prevalence (40%) of UTI. Studies showed that people with little or no formal education had higher prevalence of UTIs ^[30,31]^. However, this report contradicts the present findings. Similar to the results of this study is that of Ndako *et al*. ^[19]^ and Mokube *et al*. ^[8]^

On the basis of occupation, UTI was higher among students (38.5%). This corroborated previous finding ^[32,33]^. The χ^2^ test of participants positive for UTI showed statistical significant (*P*<.05) with respect to occupation (*P*=.003). No significant variations (*P*□.05) was observed with respect to age (*P*=.946), sex (*P*=.918), marital status (*P*=.347) and educational level (*P*=.508) (Table 2).

Of all the clinical symptoms, dysuria was significantly associated with UTI (*P*=.033, [95%CI; 1.051-4.059]). This contradicted Seifu and Gebissa ^[34]^ who reported that fever, urgency, frequency and supra-pubic pain were significantly associated with UTI, but corroborated Al-Kashif ^[35]^ who reported statistical association of dysuria with UTI.

In line with earlier studies ^[5,6,35,36]^ pregnancy (*P*=.001, [95%CI; 1.858-12.575]), previous history of UTI (*P*=.000, [95%CI; 1.582-5.180]), history of catheterization (*P*= .031, [95%CI; 1.024-19.053]) and use of antibiotics without prescription (*P*=.000, [95%CI; 0.040-0.220]) were found significantly associated with the occurrence of UTI. This may be attributed to factors such as; resistant strains of uropathogens from the earlier episodes, contamination catheter during insertion, and physiological changes. More so, contrary to Odoki *et al*. ^[10]^ and Ahmed ^[37]^ the present study revealed that, use of contraceptive (*P*=.296, [95%CI; 0.658-3.900]), family history of UTI (*P*=.489, [95%CI; 0.687-2.191]) and diabetes (*P*=.142, [95%CI; 0.455-57.368]) had no significant association with UTI.

The severity of UTI is greatly influenced by the types of organisms involved. In this study, Gram negative uropathogens constituted 80% and Gram positive (20%). This supported the assertion that Gram negative bacteria constituted 80-90% of uropathogens ^[38]^. The uropathogens recovered in this study had previously known to cause UTI ^[6,7,17,26]^.

The highest prevalence of *K. pneumoniae* in this study is an indication that the organism is achieving more prominence as causative agents of UTI. This finding is inconsistence with Seifu and Gebissa ^[34]^ in Ethiopia who reported *E. coli* as the commonly uropathogens from UTI patients. In other reports, Many *et al*. ^[39]^ in DRC, reported *P. mirabilis* (41.2%), Labi *et al*.^[24]^ in Ghana, reported *Enterococcus* sp (26.7%) and Musonda *et al*. ^[40]^ in Zambia, reported *Staphylococcus aureus* (32%) as the most predominant organism. The variation in the type of bacteria uropathogens in this study and others reported might be attributed to physiological state of patients, techniques of sample collection, sample size used, environmental or personal hygiene.

This study revealed the presence of *Cronobacter* sp in the analyzed urine samples. *Cronobacter* sp had been reported as emerging pathogens from infant food ^[41]^. The incidence of UTI cause by *Cronobacter* sp in the study area is rear. However, this study corroborated with Hayashi *et al*. ^[42]^ who reported the occurrence of *C. sakazakii* in a 69 years old man presenting with UTI in Shimane University Hospital, Shimane, Japan. The possible rout of infection with *Cronobacter* sp could be via oral ingestion from external sources and retrograde UTI ^[42]^.

## CONCLUSION

The overall prevalence of UTI among patients attending hospitals in Calabar, Nigeria was 28.6%. Of all considered risk factors, previous history of UTI, use of drugs without prescription, pregnancy and history of urinary catheter were significantly (*P*<.05) associated with the occurrence of UTI. Similarly, there was association of dysuria and occupation with the occurrence of UTI. *K. pneumoniae* was the most predominant uropathogens, followed by Coagulase-negative Staphylococci. Other isolates were *E. coli, E. clocae, C. freundii, P. mirabilis, S. marcescens, P. aeruginosa, Cronobacter* sp, *Enterococcus* sp, *C. koseri*, and *P. luteola*. Therefore, routine UTI screening is recommended for pregnant women, patients with difficult or painful urination, patients with previous episodes of UTI, catheterized patients and appropriate drugs administered for positive cases. In addition, self-administration of antibiotics should be avoided.

## Supporting information

Questionnaire and identification table

## Data Availability

Data regenerated in this study are available upon request by the publisher or any researcher provided it is not for commercial purposes.

## ACKNOWLEDGEMENTS

Authors would like to appreciate the participants, management and staff of the University of Calabar Teaching Hospital (UCTH), Nigeria Navy Reference Hospital Calabar (NNRH), and General Hospital Calabar (GHC), for their contributions towards the success of this research.

