## Supplementary material for "Prevalence of Symptomatic Bacteriuria and Associated Risk Factors among Patients Attending Major Hospitals in Calabar, Nigeria": Questionnaire and identification table

**Appendix A1: Sample Questionnaire:**

**SECTION (A): Socio-Demographic Data**

1. **Age in years**: What is your age range: 5-18 { } 18-29 { } 30-45 { } ≥ 45{ }

2. **Sex**: Male { } Female { }

3. **Marital status**: Single { } Married { } Divorce { } Widow { }

4. **Highest educational qualification**: No formal education { } Primary { }

Secondary { } Tertiary { } Others (please specify) ……………….

5. **Major occupation**: Farming { } Fishing { } Trading { } Student { } Civil servant { } Artisan { } others; specify ……………………………….

**SECTION (B): Risk Factors**

6. **Pregnancy status** (for women only): Pregnant { } Not pregnant { }

7. **Antibiotics usage**: When last did you take antibiotics? Currently on antibiotics { }< one week { }> one week but < two weeks { }> two weeks { }

8. **History of hospitalization**: One week { } two weeks { } three weeks { } four weeks { }

more than four weeks { }

9. **Use of antibiotics**: Did you use antibiotics without prescription? Yes { } No { }

10. **Previous history of UTI**: Have you had urinary tract infection before? Yes { } No { }

11. **History of catheterization**: Did you use urinary catheter before? Yes { } No { }

12 **Family histories**: Has any member of your family previously diagnosed of UTI? Yes {	}

No { }

13. **Diabetes**: Are you diabetic? Yes { } No { }

14. **Frequency of urination**: How often do you urinate in an hour? Once { } 2-4 times { }

5-8 times { } > than 8 times { }

15. **Use of contraceptive**: Have you been using any contraceptive? Yes { } No { }

16. **Urine urgency**: Do you feel like urinating on yourself if you don’t quickly go to urinate? Yes { } No { }

17. **Dysuria**: Do you have pain or difficulty during urination? Yes { } No { }

18. **Fever**: Do you have high body temperature? Yes { } No { }

19. **Flank or supra-pubic pain**: Do you have back or abdominal pain? Yes { } No { }

**Appendix A2: Identification of Gram negative uropathogens using API 20E**

| **ISOLATE CODE** | **ONPG** | **ADH** | **LDC** | **ODC** | **CIT** | **H_2_S** | **URE** | **TDA** | **IND** | **VP** | **GEL** | **GLU** | **MAN** | **INO** | **SOR** | **RHA** | **SAC** | **MEL** | **AMY** | **ARA** | **OX** | **PROFILE CODE** | **% ID** |
| --- | --- | --- | --- | --- | --- | --- | --- | --- | --- | --- | --- | --- | --- | --- | --- | --- | --- | --- | --- | --- | --- | --- | --- |
| NH 59*_Cronobacter_* _sp_ | + | + | - | + | + | - | - | - | + | + | - | + | + | + | - | + | + | + | + | + | - | 3345373 | 99.9 |
| NH 4*_P. mirabilis_* | - | - | - | + | - | + | + | + | - | - | + | + | - | - | - | - | - | - | - | - | - | 0536000 | 99.9 |
| NH 3*_K. pneumoniae_* | + | - | + | - | + | - | + | - | - | + | - | + | + | + | + | + | + | + | + | + | - | 5215773 | 97.3 |
| NH 25*_E. clocae_* | + | + | - | + | + | - | - | - | - | + | - | + | + | + | + | + | + | + | + | + | - | 3305773 | 91.4 |
| UC 19*_E. coli_* | + | - | + | + | - | - | - | - | + | - | - | + | + | - | + | + | + | + | - | + | - | 5144572 | 99.5 |
| NH 17*_S. marcessens_* | + | - | + | + | + | - | - | - | - | + | + | + | + | + | + | - | + | + | + | - | - | 5307761 | 97.3 |
| GH 61*_E. coli_* | + | - | + | + | - | - | - | - | + | - | - | + | + | - | + | + | - | + | - | + | - | 5144552 | 99.9 |
| NH 10*_C. freundii_* | + | + | - | - | + | + | - | - | - | - | - | + | + | + | + | + | + | + | + | + | - | 3604773 | 99.9 |
| GH 14*_S. marcescens_* | + | - | + | + | + | - | - | - | - | + | + | + | + | + | + | - | + | + | + | - | - | 5307761 | 97.3 |
| GH 10*_C. koseri_* | + | + | - | + | + | - | - | - | + | - | - | + | + | - | + | + | - | - | + | + | - | 3344513 | 99.9 |
| GH 15*_P. mirabilis_* | - | - | - | + | - | + | + | + | - | - | + | + | - | - | - | - | - | - | - | - | - | 0536000 | 99.9 |
| UC 31*_E. coli_* | + | - | + | + | - | - | - | - | + | - | - | + | + | - | + | + | - | - | - | + | - | 5144512 | 98.1 |
| UC 41*_K. pneumoniae_* | + | - | + | - | - | - | - | - | - | + | - | + | + | + | + | + | + | + | + | + | - | 5005773 | 96.0 |
| NH 22*_C. freundii_* | + | + | - | - | + | + | - | - | - | - | - | + | + | - | + | + | + | + | - | + | - | 3604572 | 99.9 |
| NH 5*_E. coli_* | - | - | + | - | - | - | - | - | + | - | - | + | + | - | + | + | - | - | - | + | - | 4044512 | 99.8 |
| GH 50*_Cronobacter_* _sp_ | + | + | - | + | + | - | - | - | - | + | - | + | + | + | - | + | + | + | + | + | - | 3305373 | 98.5 |
| GH 67*_P. mirabilis_* | - | - | - | + | + | + | + | + | - | - | + | + | - | - | - | - | - | - | - | - | - | 0736000 | 99.9 |
| UC 40*_S. marcescens_* | + | - | + | + | + | - | + | - | - | + | + | + | + | + | + | - | + | + | + | + | - | 5317763 | 88.5 |
| UC 20*_C. freundii_* | + | + | - | - | + | + | - | - | - | - | - | + | + | - | + | + | + | + | + | + | - | 3604573 | 99.8 |
| NH 85*_E. coli_* | + | - | + | + | - | - | - | - | + | - | - | + | + | - | + | + | + | + | - | + | - | 5144572 | 99.5 |
| GH 25*_E. clocae_* | + | + | - | + | + | - | - | - | - | + | - | + | + | - | + | + | + | + | + | + | - | 3305573 | 97.7 |
| GH 8*_E. coli_* | + | - | + | + | - | - | - | - | + | - | - | + | + | - | + | + | + | + | - | + | - | 5144572 | 99.5 |
| UC 48*_K. pneumoniae_* | + | - | + | - | + | - | - | - | - | + | - | + | + | + | + | + | + | + | + | + | - | 5205773 | 81.8 |
| GH 63*_K. pneumoniae_* | + | - | + | - | + | - | + | - | - | + | - | + | + | + | + | + | + | + | + | + | - | 5215773 | 97.3 |
| GH 3*_E. coli_* | - | - | + | - | - | - | - | - | + | - | - | + | + | - | - | - | - | - | - | + | - | 4044102 | 97.3 |
| GH 7*_C. freundii_* | + | - | - | - | + | + | - | - | - | - | - | + | + | - | + | + | + | + | + | + | - | 1604573 | 99.8 |
| UC 13*_K. pneumoniae_* | + | - | + | - | - | - | - | - | - | - | - | + | + | + | + | + | + | + | + | + | - | 5004773 | 89.8 |
| NH 2*_E. clocae_* | + | + | - | + | + | - | - | - | - | + | - | + | + | - | + | - | + | + | + | + | - | 3305563 | 99.3 |

**Appendix A2: Continue**

| **ISOLATE CODE** | **ONPG** | **ADH** | **LDC** | **ODC** | **CIT** | **H_2_S** | **URE** | **TDA** | **IND** | **VP** | **GEL** | **GLU** | **MAN** | **INO** | **SOR** | **RHA** | **SAC** | **MEL** | **AMY** | **ARA** | **OX** | **PROFILE CODE** | **% ID** |
| --- | --- | --- | --- | --- | --- | --- | --- | --- | --- | --- | --- | --- | --- | --- | --- | --- | --- | --- | --- | --- | --- | --- | --- |
| NH 26*_P. mirabilis_* | - | - | - | + | - | - | + | + | - | - | + | + | - | - | - | - | - | - | - | - | - | 0136000 | 99.9 |
| NH 28*_P. mirabilis_* | - | - | - | + | + | + | + | + | - | - | - | + | - | - | - | - | - | - | - | - | - | 0734000 | 99.9 |
| NH 69*_K. pneumoniae_* | + | - | + | - | - | - | - | - | - | + | - | + | + | + | + | + | + | + | + | + | - | 5005773 | 96.0 |
| UC 33*_K. pneumoniae_* | + | - | + | - | + | - | - | - | - | + | - | + | + | + | + | + | + | + | + | + | - | 5205773 | 81.8 |
| NH 78*_K. pneumoniae_* | + | - | - | - | - | - | + | - | - | + | - | + | + | + | + | + | + | + | + | + | - | 1015773 | 98.0 |
| GH 1*_S. marcescens_* | + | - | + | + | + | - | + | - | - | + | + | + | + | + | + | - | + | + | + | - | - | 5317761 | 99.8 |
| UC 25*_K. pneumoniae_* | + | - | + | - | + | - | + | - | - | + | - | + | + | + | + | + | + | + | + | + | - | 5215773 | 97.3 |
| GH 11*_C. freundii_* | + | + | - | - | + | + | - | - | - | - | - | + | + | + | + | + | + | + | + | + | - | 3604773 | 99.9 |
| GH 37*_K. pneumoniae_* | + | - | + | - | + | - | - | - | - | - | - | + | + | + | + | + | + | + | + | + | - | 5204773 | 81.1 |
| GH 23*_C. freundii_* | + | + | - | - | + | + | - | - | - | - | - | + | + | - | + | + | + | + | - | + | - | 3604572 | 99.9 |
| NH 34*_E. clocae_* | + | + | - | + | + | - | - | - | - | - | - | + | + | - | + | - | + | + | + | + | - | 3304563 | 87.4 |
| NH 23*_S. marcescens_* | + | - | + | + | + | - | + | - | - | + | + | + | + | + | + | - | + | + | + | + | - | 5317763 | 88.5 |
| NH 66*_K. pneumoniae_* | + | - | + | - | - | - | + | - | - | + | - | + | + | + | + | + | + | + | + | + | - | 5015773 | 94.0 |
| NH 32*_K. pneumonia_* | + | + | + | - | - | - | + | - | - | + | - | + | + | + | + | + | + | + | + | + | - | 7015773 | 95.1 |
| NH 33*_K. pneumoniae_* | + | - | - | - | - | - | + | - | - | + | - | + | + | + | + | + | + | + | + | + | - | 1015773 | 98.0 |
| NH 29*_E. clocae_* | + | - | - | + | + | - | - | - | - | + |  | + | + | - | + | + | + | + | + | + | - | 1305573 | 93.4 |
| NH 30*_E. coli_* | - | - | + | + | - | - | - | - | + | - | - | + | + | - | + | + | - | - | - | + | - | 4144512 | 69.5 |
| NH 74*_K. pneumoniae_* | + | - | + | - | - | - | - | - | - | - | - | + | + | + | + | + | + | + | + | + | - | 5004773 | 89.8 |
| UC 32*_E. clocae_* | + | + | - | + | + | - | - | - | - | - | - | + | + | - | + | - | + | + | + | + | - | 3304563 | 87.4 |
| GH 9*_E. clocae_* | + | + | - | + | + | - | - | - | - | + | - | + | + | - | + | + | + | + | + | + | - | 3305573 | 97.7 |
| NH 63*_K. pneumoniae_* | + | - | + | - | - | - | - | - | - | + | - | + | + | + | + | + | + | + | + | + | - | 5005773 | 96.0 |

**KEY:**

NH=Isolates from Nigeria Navy Reference Hospital Calabar, UC=Isolates from University of Calabar Teaching Hospital, GH=Isolates from General Hospital Calabar. ONPG=Ortho-Nitrophenyl-β-galactosidase, TDA=Triptophan deaminase, H_2_S=Hydrogen sulphide, VP=Voges-Proskauer, ADH=Arginine dihydrolase, LDC=Lysine decarboxylase, ODC=Ornithine decarboxylase, CIT=Citrate utilization. URE=Urease, IND=Indole production, GEL=Gelatinase, GLU=Glucose, MAN=Mannitol, INO=Inositol, SOR=Sorbitol, RHA=Rhamnose, SAC=Sucrose, MEL=Melibiose, AMY=Amygdain, ARA=Arabinose, OX=Oxidase, %ID=Percentage probability.

**Appendix A3: Identification of Gram negative uropathogens using API 20NE**

| **ISOLATE CODE** | **NO_2_** | **TRP** | **GLU** | **ADH** | **URE** | **ESC** | **GEL** | **PNG** | **GLU** | **ARA** | **MNE** | **MAN** | **NAG** | **MAL** | **GNT** | **CAP** | **ADI** | **MLT** | **CIT** | **PAC** | **OX** | **PROFILE CODE** | **% ID** |
| --- | --- | --- | --- | --- | --- | --- | --- | --- | --- | --- | --- | --- | --- | --- | --- | --- | --- | --- | --- | --- | --- | --- | --- |
| NH 20*_P. luteola_* | + | - | - | + | - | + | - | + | + | + | + | + | - | + | + | + | - | + | + | - | - | 1567651 | 99.9 |
| NH 15*_P. aeruginosa_* | + | - | - | + | - | - | + | - | + | - | - | + | + | - | + | + | + | + | + | - | + | 1154575 | 99.5 |
| UC 23*_P. aeruginosa_* | + | - | - | + | + | - | + | - | + | - | - | + | + | - | + | + | + | + | + | - | + | 1354575 | 99.9 |

KEY:

NH=Isolates from Nigeria Navy Reference Hospital Calabar, UC=Isolates from University of Calabar Teaching Hospital, NO2=Potassium nitrate, TRP=Indole production, GLU=Glucose, ADH=Arginine dihydrolase, URE=Urease, ESC=Esculin hydrolysis, GEL=Gelatin hydrolysis, PNPG=β-galactosidase, ARA=Arabinos, MNE=Mannose, MAN=Mannitol, NAG=N-Acetyl-Glucosamine, MAL=Maltose, GNT=Potassium gluconate, CAP=Capric acid, ADI=Adipic acid, MLT=Malic acid, CIT=Trisodium citrate, PAC= Phenylacetic acid, OX= Oxidase, %ID= Percentage probability.
